# Novel surrogate virus neutralization test reveals low serum neutralizing anti-SARS-CoV-2-S antibodies levels in mildly affected COVID-19 convalescents

**DOI:** 10.1101/2020.07.12.20151407

**Authors:** Berislav Bošnjak, Saskia Catherina Stein, Stefanie Willenzon, Anne Katrin Cordes, Wolfram Puppe, Günter Bernhardt, Inga Ravens, Christiane Ritter, Christian R. Schultze-Florey, Nina Gödecke, Jörg Martens, Hannah Kleine-Weber, Markus Hoffmann, Anne Cossmann, Mustafa Yilmaz, Isabelle Pink, Marius M. Hoeper, Georg M.N. Behrens, Stefan Pöhlmann, Rainer Blasczyk, Thomas F. Schulz, Reinhold Förster

## Abstract

Neutralizing antibodies targeting the receptor-binding domain (RBD) of the SARS-CoV-2 spike (S) block severe acute respiratory syndrome coronavirus 2 (SARS-CoV-2) entry into cells using surface-expressed angiotensin-converting enzyme 2 (ACE2). We developed a surrogate neutralization test (sVNT) to assess at what degree serum antibodies interfere with the binding of SARS-CoV-2-S-RBD to ACE2. The sVNT revealed neutralizing anti-SARS-CoV-2-S antibodies in the sera of 90% of mildly and 100% of severely affected coronavirus-disease-2019 (COVID-19) convalescent patients. Importantly, sVNT results correlated strongly to the results from pseudotyped-vesicular stomatitis virus-vector-based neutralization assay and to levels of anti-SARS-CoV-2-S1 IgG and IgA antibodies. Moreover, levels of neutralizing antibodies also correlated to duration and severity of clinical symptoms, but not patient age or gender. These findings together with the sVNT will not only be important for evaluating the prevalence of neutralizing antibodies in a population but also for identifying promising plasma donors for successful passive antibody therapy.

## Introduction

Within 6 months since its emergence, the novel severe acute respiratory syndrome coronavirus 2 (SARS-CoV-2), the cause of coronavirus disease 2019 (COVID-19), has rapidly spread around the globe. COVID-19 consists of a spectrum of clinical syndromes ranging from asymptomatic cases through mild, flu-like disease to severe illness requiring hospitalization mainly due to pulmonary complications (*1–3*). Although SARS-CoV-2 primarily targets the respiratory system, new data indicate that COVID-19 also affects the vascular system, causing thrombotic microangiopathy and thrombosis in multiple organs, including the lungs (*4–6*). It is not surprising, therefore, that patients with pre-existing cardiovascular diseases, hypertension, and other co-morbidities are particularly endangered (*7*).

SARS-CoV-2 uses angiotensin-converting enzyme 2 (ACE2) as receptor for entry into target cells and employs TMPRSS2, a cellular serine protease for activation of the viral spike (S) protein (*8, 9*). Both ACE2 and TMPRSS2 are particularly abundant in the upper respiratory tract (*10*). An early immune response against SARS-CoV-2 involves interleukin (IL)-6 and interferon-signature gene expression in alveolar macrophages and infiltrating monocytes (*11*). Although this early immune response is accompanied by severe lymphopenia (*12, 13*), a growing amount of data indicate that successful recovery from COVID-19 relies on antibody and T cell responses (*12, 14–17*). Importantly, there appears to be a strong correlation between circulating SARS-CoV-2-specific CD4^+^ and CD8^+^ T cells and IgG antibodies against the nuclear (N) and/or the spike (S) protein of SARS-Cov-2 (*16, 17*).

Current data indicate that anti-SARS-CoV-2 IgM antibodies appear within one week after infection, remain present for a month before they gradually decrease (*18, 19*). In contrast, anti-SARS-CoV-2 IgG antibodies appear within 10-21 days after infection and seem to remain more-or-less stable for up to 3 months (*18, 19*). It is not surprising therefore, that antibody responses against SARS-CoV-2 have also received attention as a method for accurate assessment of infection prevalence (*20, 21*). Particularly interesting are antibodies targeting the receptor-binding domain (RBD) of the S protein, as they can block virus entry into the cells and thus prevent infection and spread. Furthermore, these neutralizing antibodies could also be used for passive antibody therapy (*20, 22*).

The field is rapidly evolving and there is still no consensus about the diagnostic value of divergent ELISA-based antibody tests for SARS-CoV-2 seropositivity (*23*). Moreover, there is uncertainty about the durability of anti-SARS-CoV-2 antibody-responses, especially as there are indications that antibody responses to other coronaviruses are variable and transient (*24–27*). It is also not clear whether all COVID-19 patients, especially those with mild disease, will raise sufficient amounts of neutralizing antibodies against SARS-CoV-2, to prevent early re-infection. In the present study we applied different experimental approaches to qualitatively and quantitatively assess the antibody response to SARS-CoV-2 infection primarily in cohorts of convalescent individuals with mild COVID-19 disease. We developed an ELISA-based surrogate virus neutralization test (sVNT) detecting antibodies that interfere with the binding of SARS-CoV-2-S-RBD protein to ACE2 and compared it to an established pseudotyped-vesicular stomatitis virus (VSV)-vector-based neutralization assay (pVNT). Furthermore, we correlated the functional data to anti-SARS-CoV-2-S1 IgG and IgA levels measured by a commercial S1 protein ELISA as well as to clinical parameters. We found low titers of neutralizing anti-SARS-CoV-2-S antibodies in 93% of convalescent patients with mild COVID-19. Importantly, only low levels of neutralizing anti-SARS-CoV-2-S titers could be detected in those convalescents that exhibited clinical symptoms for a short period of time. In contrast, sera from convalescents with severe COVID-19 contained significantly higher total and neutralizing antibody titers. The newly developed sVNT allows for high throughput screening, which would be valuable for epidemiological studies as well as for identifying suitable plasma donors for passive immunization.

## Results

### Most individuals with mild COVID-19 disease develop anti-SARS-CoV-2 antibodies

Between March 23^rd^ and May 11^th^ 2020, we enrolled in a first step a total of 51 convalescent COVID-19 individuals diagnosed with SARS-CoV-2 infection by RT-PCR and 12 healthy control subjects that were not exposed to SARS-CoV-2. The samples from convalescent COVID-19 cases were split into two groups according to disease severity. 80% of the individuals (n = 40) had a mild clinical course with average symptom duration of 10 days (range, 0-25 days) which did not require an in-patient hospital stay (Tables S1 and S2). Ten patients had severe COVID-19 of who required hospital stays longer than 2 weeks and respiratory support. Severe COVID-19 patients had average disease duration of 37 days (range from 19-71 days).

To estimate the overall antibody responses against SARS-CoV-2 in the serum of convalescent COVID-19 individuals, we analyzed the presence of anti-SARS-CoV-2 IgG and IgA antibodies targeting the S1 protein by ELISA. Anti-SARS-CoV-2-S1 specific IgG antibodies were detected in 35/37 (94.6%) of the mildly affected and in 10/10 severely affected COVID-19 patients tested. One individual with mild disease was considered to have borderline serum positivity while one patient was negative according to the manufacturer’s classification (Fig. 1A). Similarly, anti-SARS-CoV-2-S IgA antibodies were present in 33/37 (89.2%) tested sera; two were diagnosed as borderline positive and two as negative (Fig. 1B). All sera from tested healthy controls (8/8) were negative for SARS-CoV-2-S-specific IgG and IgA (Fig. 1, A and B).

**Fig 1.**
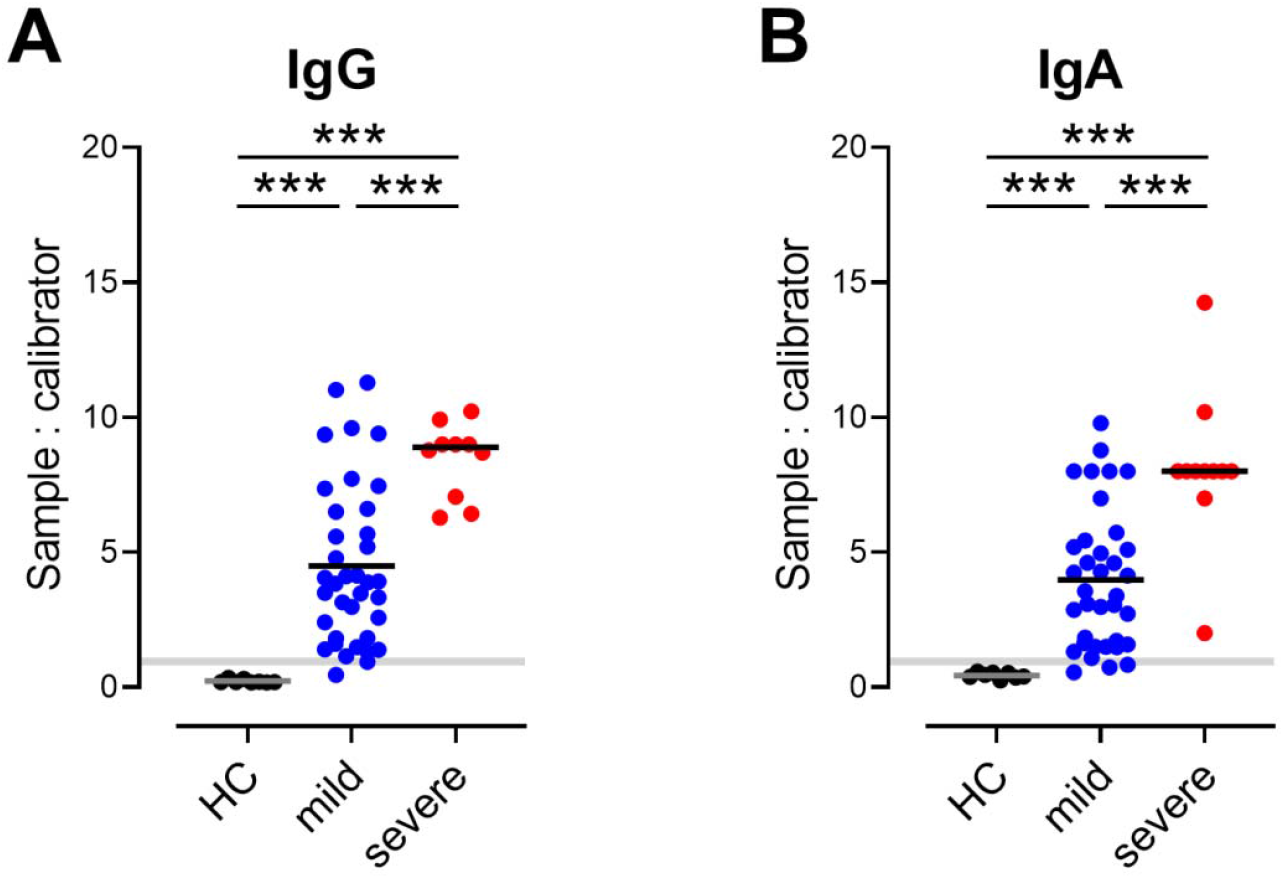
Qualitative analysis of serum total IgG (A) and IgA (B) antibodies against SARS-CoV-2 S1 in convalescent patients with mild or severe COVID-19 and healthy controls (HC) determined by ELISA. Shaded areas cut-off values to determine positive (above), borderline (within) and negative (below gray area) samples. Dots, individuals; bars, mean, *** P < 0.001, Welch’s ANOVA followed by Dunnett’s T3 multiple comparisons test.

### A surrogate virus-neutralization assay for the detection of neutralizing antibodies

Based on the consideration that serum neutralizing antibodies might also reduce the binding of SARS-CoV-2 to ACE2 *in vitro*, we developed a surrogate virus-neutralization assay. At the outset, we coated microtiter plates with different amounts of ACE2, obtained from a commercial vendor or produced in-house and titrated soluble His-tagged SARS-CoV-2-S-RBD. Binding was revealed by a peroxidase-labeled anti-His mAb and a chromogenic substrate (Fig. 2, A and B). The dissociation constant (K_D_) in our assay was 5.9±1.9 nM (SD±SEM) and thus similar to values reported elsewhere (*28*).

**Fig 2.**
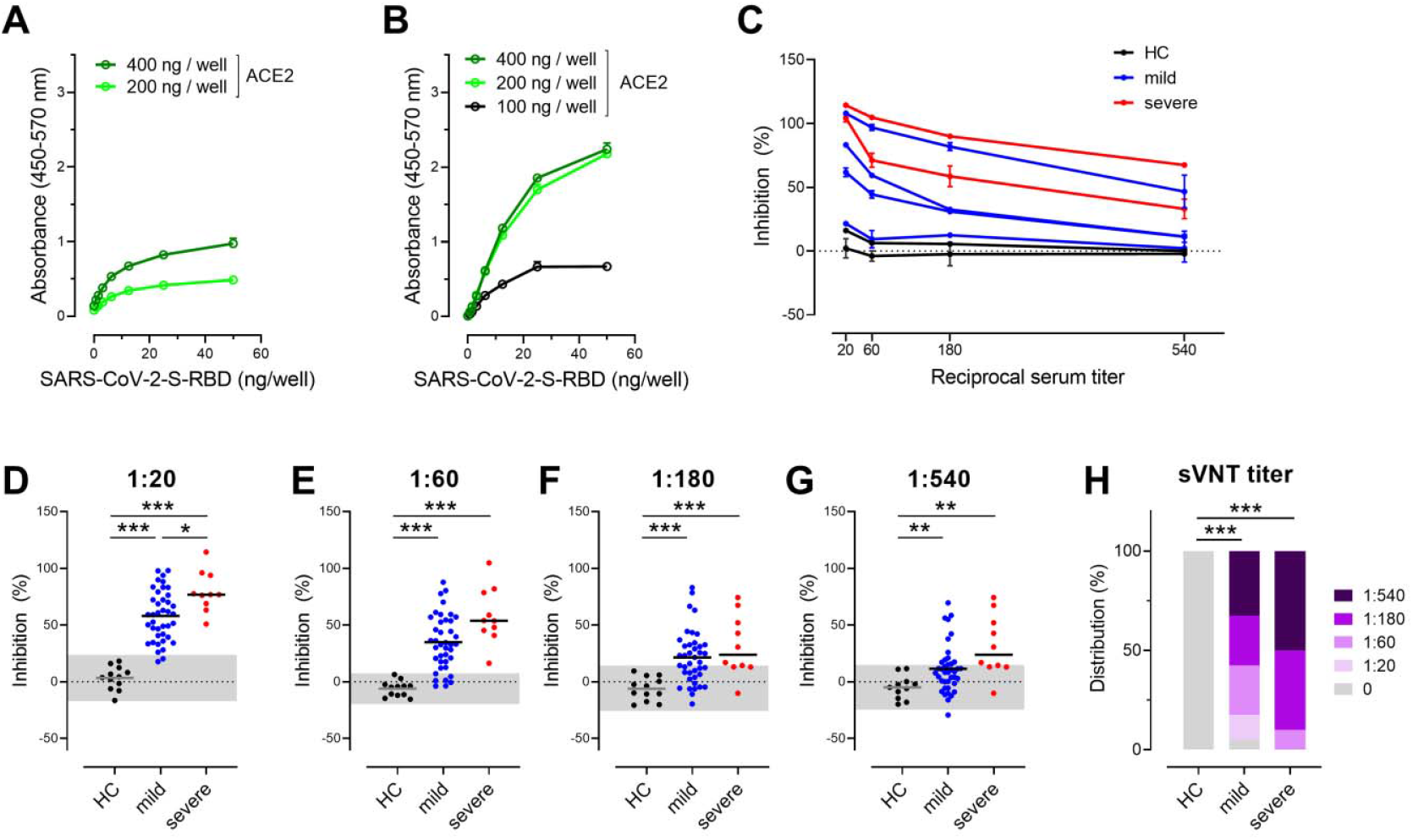
Surrogate virus neutralization test (sVNT) detects neutralizing antibodies interfering with SARS-CoV-2-S-RBD binding to human ACE2. (A,B) Binding of SARS-CoV-2-S-RBD to human ACE2 from commercial vendor (A) and produced in-house (B). Plates were coated with ACE2 as indicated. His-tagged SARS-CoV-2-S-RBD was titrated as indicated and revealed with anti-His peroxidase-labeled mAb. Representative assays performed in duplicates, mean ± SD. (C) Inhibition of interaction of SARS-CoV-2-S-RBD with ACE2 by the addition of sera of convalescent patients with mild (blue lines) or severe (red lines) COVID-19 and healthy controls (HC; black lines). Assay performed in duplicates; mean percentages of neutralization ± SD. (D-G) Inhibition of interaction of SARS-CoV-2-S-RBD with ACE2 at serum dilutions indicated. Individual values (dots) and means (line). Shaded areas represent mean ± 2 SD of values from sera of healthy controls. * P < 0.05; *** P < 0.001, Welch’s ANOVA followed by Dunnett’s T3 multiple comparisons test. (H) Relative distributions of SARS-CoV-2 neutralizing serum titers determined as dilution that still had binding reduction > mean + 2 SD of HC. *** P < 0.001; Fisher’s exact test (HC *vs*. mild or severe) or Chi-squared test for trend (mild *vs*. severe).

Based on these findings, we used 300 ng/well of commercially or 150 ng/well of in-house produced ACE2 in combination with 6 ng/well SARS-CoV-2-S-RBD-His to test sera for their abilities to interfere with the binding of these two proteins. Indeed, sera from convalescents could interfere with S protein binding to ACE2, although with different efficiencies, while sera from healthy controls did not (Fig. 2C). Sera from convalescent’s were scored ‘positive’ at any tested dilution once binding reduction was > mean + 2 SD of values from sera of healthy controls. At serum dilutions of 1:20, this assay identified neutralizing antibodies in 38/40 (95.0%) of mildly and in 11/11 of severely affected convalescent patients but in none of the 11 tested healthy control samples (Fig. 2, C and D). With increasing serum dilutions the number of “positive” sera dropped constantly (Fig. 2, E-G). The median sVNT titer in the mildly affected convalescent cohort was 1:180 indicating that patients with mild COVID-19 develop relatively low amounts of SASRS-CoV-2 neutralizing antibodies (Fig. 2H). In contrast, severe COVID-19 convalescents had a median sVNT titer of ≥1:540 (Fig. 2H).

### Neutralizing antibodies determined by pseudotyped virus neutralization test

Next, we determined the levels of neutralizing antibodies by an established SARS-CoV-2 spike protein pseudotyped vesicular stomatitis virus (pVSV)-based neutralization test (pVNT) (*8*). This pVNT relies on the use of replication-defective VSV particles carrying SARS-CoV-2 S protein to reflect the entry of SARS-CoV-2 into host cells. Particles carrying the G-protein of VSV were used as a control. Sera from healthy controls neither suppressed VSV-G nor SARS-CoV-2-S-driven transduction. Likewise, sera from convalescents did not suppress transduction driven by VSV-G but in many cases transduction mediated by SARS-CoV-2-S was inhibited (Fig. 3A). We used this assay to determine the serum dilutions that reduced transduction of pVSV by 90% (pVNT_90_) and 50% (pVNT_50_) respectively (Fig. 3, B and C). In contrast to the 10 tested sera from severely affected COVID-19 convalescent patients that all had neutralizing antibodies, such molecules were detected in only 29 (pVNT_90_) and 37 (pVNT_50_) of the 40 tested mildly affected convalescent sera but in none of the sera from healthy controls (Fig. 3, B and C). Of note, the median pVNT_50_ in the mildly affected convalescent group was 1:100 and the median pVTN_90_ was 1:25, further supporting the finding that patients with mild COVID-19 only develop low amounts of SASRS-CoV-2 neutralizing antibodies. In contrast, the median pVNT_50_ and pVTN_90_ in severely affected COVID-19 convalescents was 1:1600 and 1:400, respectively, further indicating that the patients recovering from severe disease develop higher neutralizing anti-SARS-CoV-2 antibody titers than patients with mild disease.

**Fig 3.**
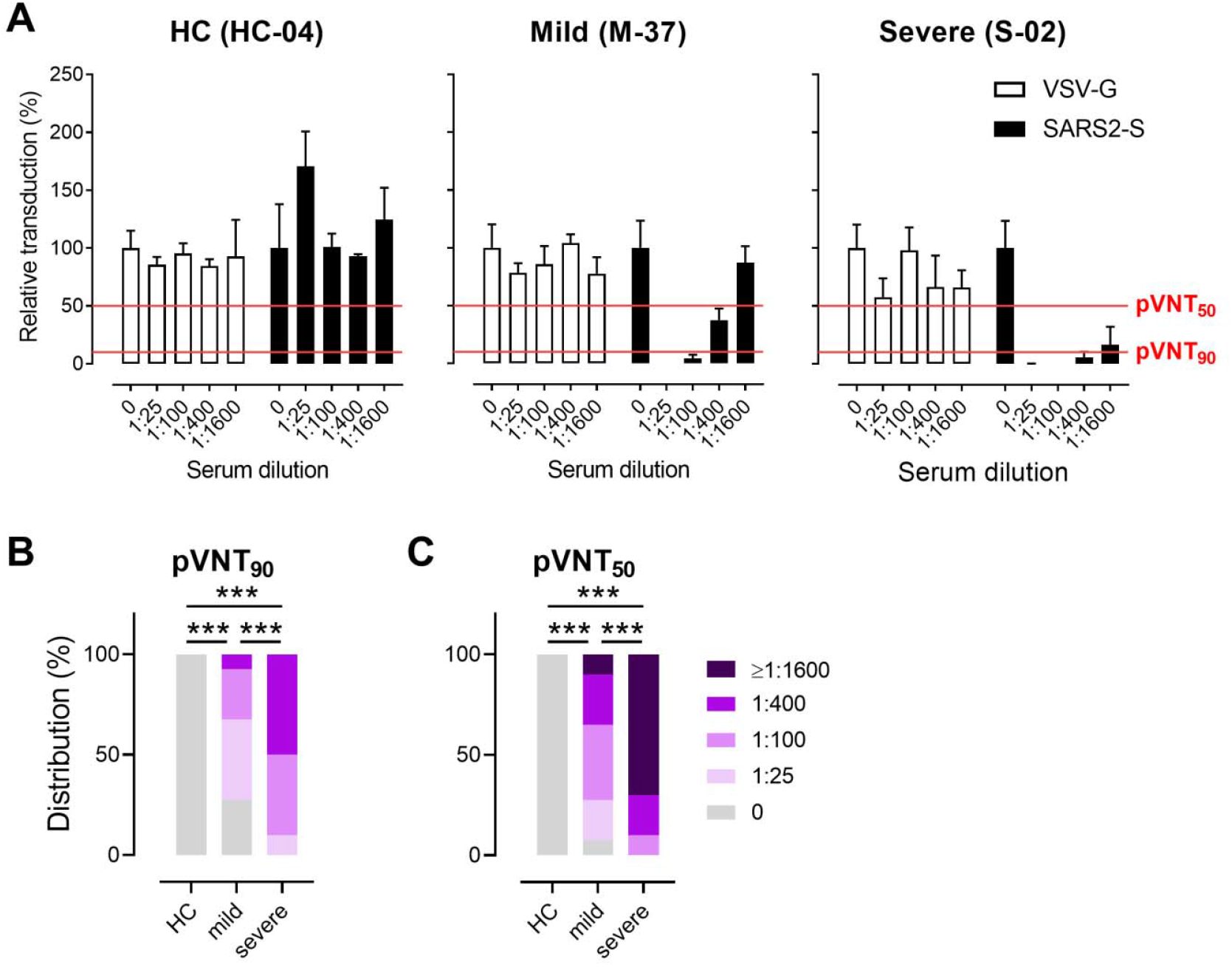
Frequency of neutralizing antibodies against SARS-CoV-2 measured by a pseudotyped virus neutralization test (pVNT) based on SARS-CoV-2 S protein pseudotyped VSV. (A) Example of pVNT results. Sera from COVID-19 convalescent patients with mild or severe disease - but not from healthy controls (HC) - suppress entry of replication-defective VSV particles carrying SARS-CoV-2 S protein into host cells (filled bars), while neither sera suppressed entry of control particles carrying the G-protein of VSV entry (open bars). Red lines indicate levels of 50% or 90% suppression of virus entry as indicated. (B,C) Relative distributions of SARS-CoV-2 neutralizing serum titers that result in (B) 90% (pVNT_90_) or (C) 50% (pVNT_50_) reduction of luciferase production as described in (A). *** P < 0.001; Fisher’s exact test (HC *vs*. mild or severe) or Chi-squared test for trend (mild *vs*. severe).

### Positive correlation between total anti-S1 protein and neutralizing antibodies levels in sera of convalescent individuals with mild COVID-19

We then analyzed the correlation between total levels of anti-S1 IgG and IgA and the amount of neutralizing antibodies in our cohort of mild COVID-19 convalescent patients and healthy controls. As expected, an initial comparison showed a strong positive correlation between levels of S protein-specific IgA and IgG antibody levels in sera (Fig. S1). More importantly, there was also a robust positive correlation between the percentage of inhibition of SARS-CoV-2-S-RBD binding to ACE2 at 1:20 serum dilution (sVNT_1:20_) and pVNT_90_ as well pVNT_50_ inhibitory titers (Fig. 4, A and B). Similarly, a strong positive correlation between sVNT_1:20_ and levels of SARS-CoV-2-S1-specific IgG an antibody levels in convalescents and healthy controls could be observed, while the levels of anti-S1 IgA showed weaker correlation (Fig. 4, C and D). These data demonstrate that the sVNT reliably detects neutralizing serum antibodies against SARS-CoV-2. Furthermore, a strong correlation with r^2^ values between 0.64 and 0.75 could be revealed when comparing SARS-CoV-2-S1-specific IgG and IgA antibody levels with pVTN_90_ and pVTN_50_ (Fig. S2).

**Fig 4.**
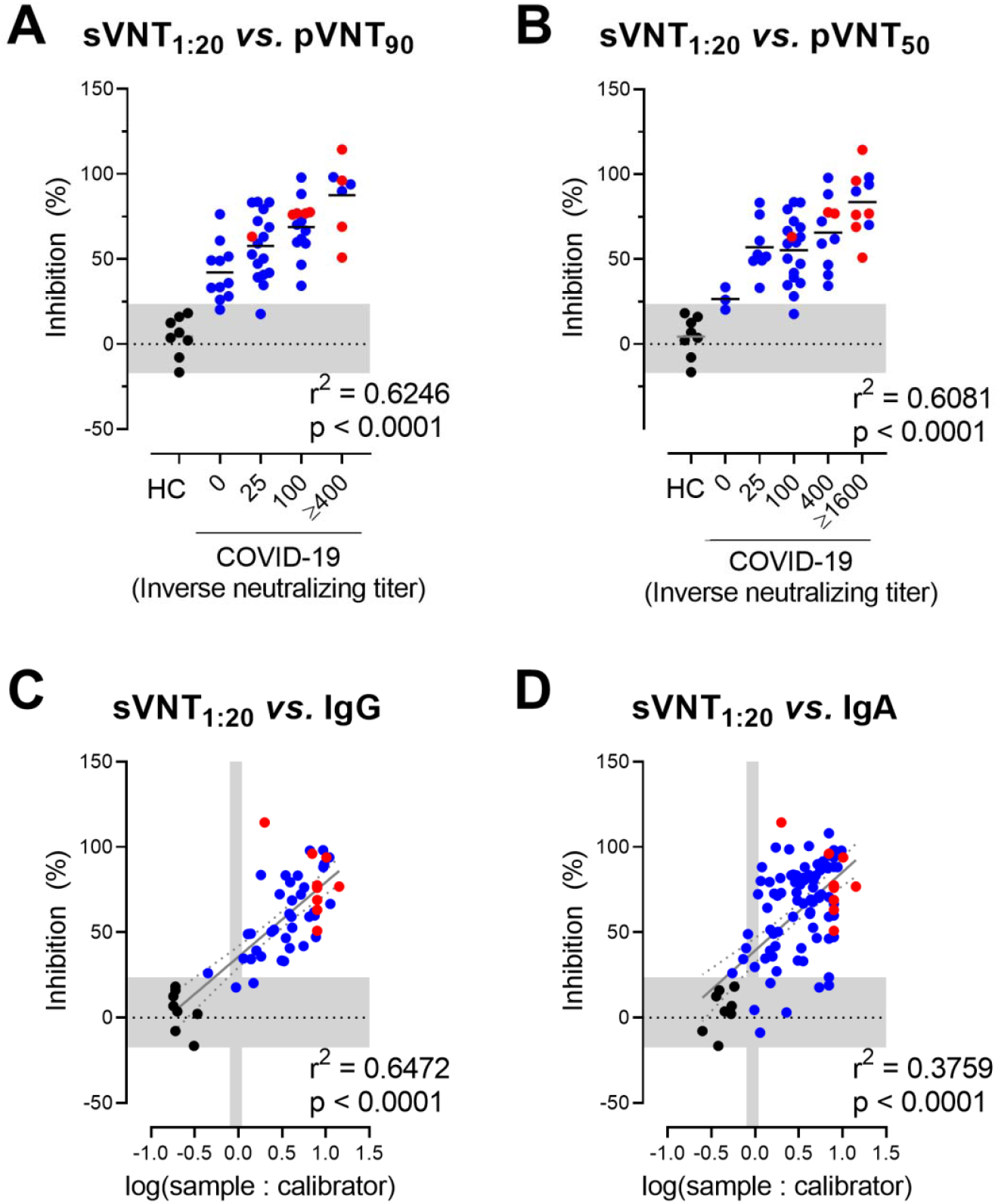
sVNT positively correlate with pVNT and anti SARS-CoV-2-S1 IgG and IgA antibodies. (A,B) Correlation between sVNT_1:20_ and antibody titers resulting in 90% (A) or 50% (B) reduction of luciferase production in pVNT_90_ and pVNT_50_. The horizontal shaded area indicates mean ± 2 S.D. range of inhibition of sera from HC. (C,D) Correlation between sVNT_1:20_ and log-transformed SARS-CoV-2-S1-specific IgG (C) and IgA (D) levels measured by ELISA. The vertical shaded areas indicate the respective cut-off values recommended by the manufacturer to determine positive (right to), borderline (within) and negative (left to) shaded area. The horizontal shaded area indicates mean ± 2 S.D. range of inhibition of sera from healthy controls. (A-D) Dots, samples of HC (black), mildly (blue) or severely (red) affected COVID-19 convalescent cases. (C,D) Correlation between the linearly-distributed data (solid line) and 95% confidence intervals (dotted lines). Correlation, one-way ANOVA followed by test for trend (A,B) or Pearson r (C,D).

### Total and neutralizing anti-SARS-CoV-2-S antibody levels in sera positively correlate with symptom duration but not timing of sampling, patient age or gender

We next examined whether the level of the protective humoral response to SARS-CoV-2 in COVID-19 patients correlated with disease duration, defined by the number of days patients showed symptoms (mild cases) or until they were discharged from the hospital (severe cases). Not surprisingly, severely affected patients had 3.5 times longer disease duration, averaging 36 days, compared to 10 days of the mildly affected patients (Fig. 5A). As severely ill patients developed higher neutralizing and total antibody titers, these data indicated that disease duration might directly influence antibody titers. This hypothesis is further supported by a positive correlation between the duration of symptoms and total anti-SARS-CoV-2 IgG, but not IgA, antibodies in convalescent patients with mild disease (Fig. 5, A and B). Although weaker, there was also a positive correlation between symptom duration and the levels of neutralizing antibodies as determined by sVNT_1:20_, pVTN_90_ and pVTN_50_ (Fig. 5, D and E). These data are in agreement with data reported from Robbiani et al (*29*) and with a recent publication indicating that asymptomatic SARS-CoV-2 infections induce lower antibody levels than symptomatic infections (*30*). Altogether, our and publicly available data suggest that a certain threshold of disease severity and/or duration might be required for successful mounting of neutralizing anti-SARS-CoV-2 humoral responses.

**Fig 5.**
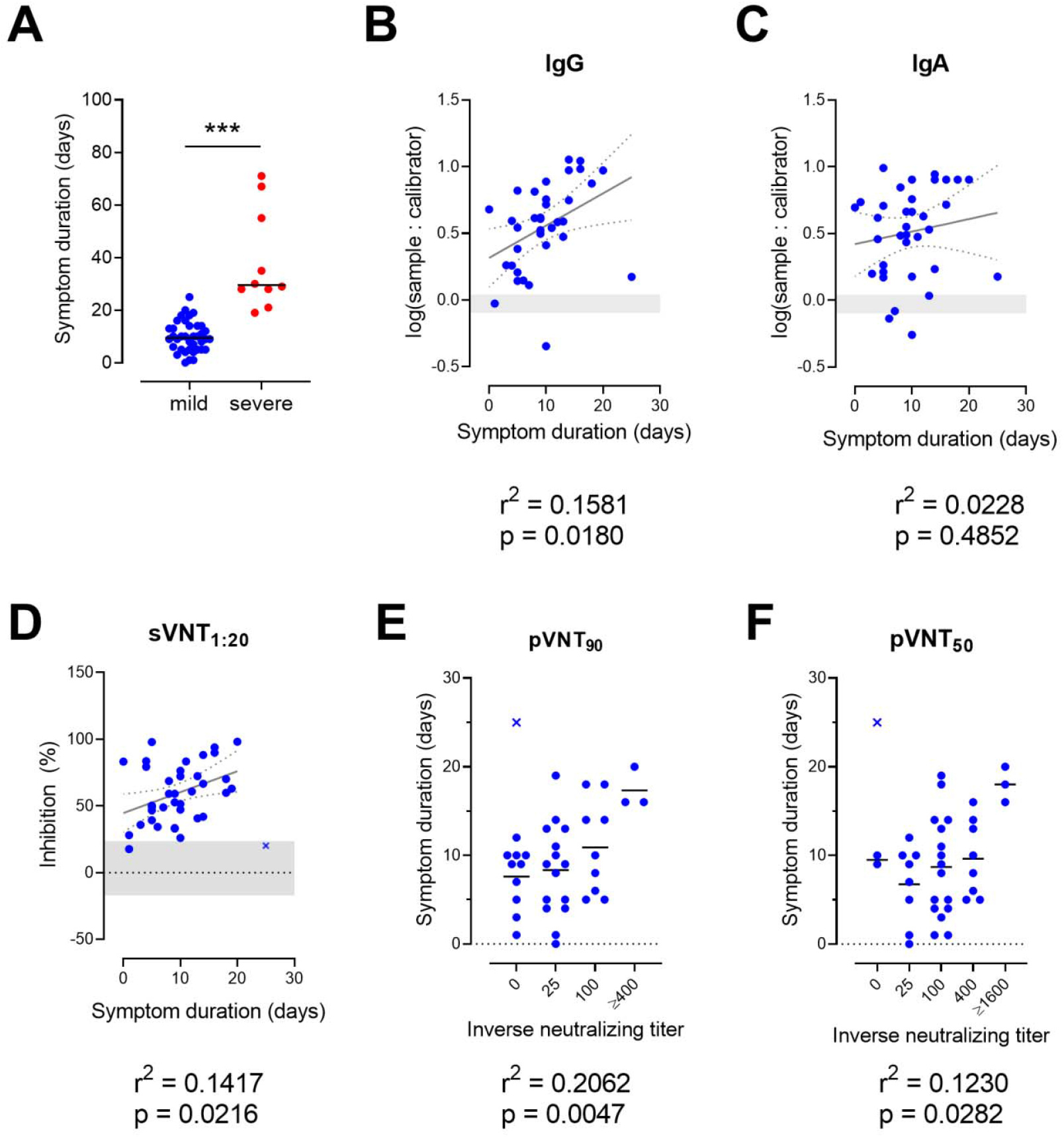
Duration of symptoms correlates to total and neutralizing SARS-CoV-2-S-specific antibody levels. (A) symptom duration of mild and severely affected COVID-19 patients. Dots, individuals; bars, mean, *** P < 0.001, Welch’s t test. (B-F) Weak positive correlation between duration of symptoms and levels of log-transformed SARS-CoV-2-S1-specific IgG (B) and IgA (C) antibodies, sVNT_1:20_ (D), pVNT_90_ (E) or pVNT_50_ (F) neutralizing antibody titers. Dots, convalescent individuals with mild COVID-19, outlier are marked with x, horizontal lines, means. (B,C) Shaded areas indicate vendor-defined cut-off values to determine positive (above), borderline (within) and negative (below gray area) samples. (D) The shaded area indicates mean ± 2 SD range of inhibition of sera from HC. Correlation, Pearson r (B-D) or one-way ANOVA followed by test for trend (E,F). Outlier was defined as a value with absolute residual value > 2SD of all residual values (D) or as a value > mean +/-2 SD of values with the same titer.

In contrast, in our cohort of convalescent COVID-19 cases, we failed to observe any correlation between levels of total and neutralizing anti-SARS-CoV-2-S antibodies with the timing of sampling after the occurrence of first symptoms (Fig. S3). Similarly, the levels of neutralizing anti-SARS-CoV-2-S IgG and IgA antibodies did not correlate with the age of patients recovered from mild disease, in spite of convalescent patients with severe disease being on average older than individuals with mild COVID-19 (Fig. S4). Of note, our cohort of convalescent COVID-19 patients with mild disease was selected based on the ability to donate blood and therefore included only 3 elderly patients (aged 60 or over). It is thus statistically underpowered to detect levels of total and neutralizing anti-SARS-CoV-2-S IgG and IgA antibodies in this age group. Males suffered more frequently from severe disease and possessed slightly increased levels of anti-SARS-CoV-2-S IgA levels compared to females (Fig. S5). However, in the cohort of recovered patients with mild COVID-19 we could not identify differences in total IgG and neutralizing anti-SARS-CoV-2-S antibodies between females and males (Fig. S5).

### sVNT allows rapid screening of sera of blood donors for the presence of neutralizing antibodies

To validate our findings, we recruited a second cohort of 44 convalescent patients with mild COVID-19 and analyzed the sera by ELISA and sVNT (Table S3). In this group of patients the ELISA detected S1 protein-specific IgA and IgG antibodies in 38 of 44 analyzed sera (borderline counted as negative; Fig. 6A). As described above for the first group of mildly affected convalescent patients, levels of S protein-specific IgA and IgG antibodies showed a strong correlation (Fig. s6). Likewise, applying the sVNT confirmed that mildly affected COVID-19 convalescent patients possess relatively low neutralizing anti-S-RBD antibodies titers (median 1:180). This assay revealed neutralizing antibodies in sera from 40 of 44 (90.9%) donors with mild COVID-19 (Fig. 6B). Along the same line, a strong positive correlation between the sVNT_1:20_ and total levels of SARS-CoV-2-S-specific IgG and IgA antibody levels could also be identified (Fig. 6, C and D). Moreover, in this group of samples, we observed a weak positive correlation between symptom duration and sVNT_1:20_ and log-transformed SARS-CoV-2-S-specific IgG and IgA levels, respectively (Fig. S7, A-C). Furthermore, significantly higher serum levels of IgA were found in males while no significant differences between males and females were found in SARS-CoV-2-S-specific IgG antibodies or in the sVNT_1:20_ (Fig. S7, D-F). As expected, we also did not find any correlation between specific IgG, IgA or sVNT_1:20_ and patient’s age or date of sampling (Fig. S7, G-I and data not shown).

**Fig 6.**
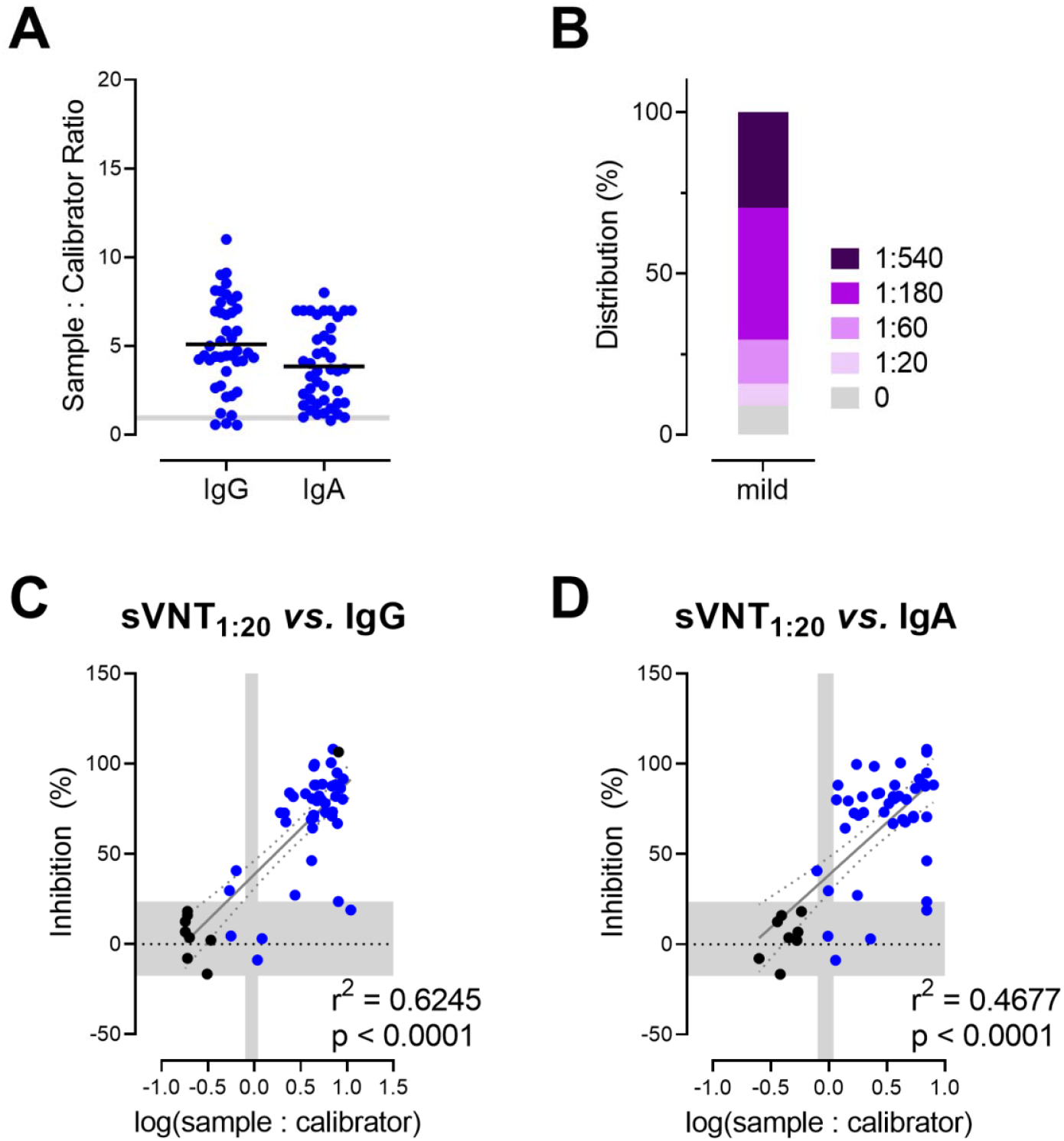
sVNT positively correlate with total levels of anti-S IgG and IgA antibodies against SARS-CoV-2-S1 in a validation cohort of convalescent patients with mild COVID-19 (n=44). (A) Serum anti SARS-CoV-2-S1 antibodies determined by ELISA. Shaded areas, cut-off values to determine positive (above), borderline (within) and negative (below gray area) samples. Dots, individuals; bars, mean. (B) Relative distribution of SARS-CoV-2 neutralizing serum titers determined as dilution that still had binding reduction > mean + 2 SD of HC. (C,D) Correlation between sVNT_1:20_ and log-transformed SARS-CoV-2-S1-specific IgG (C) and IgA (D) levels measured by ELISA. Vertical shaded areas, vendor-defined cut-off values to determine positive (right to), borderline (within) and negative (left to shaded area) samples. Horizontal shaded area, i mean ± 2 S.D. inhibition of sera from HC. Correlation, Pearson r.

Altogether, results from the validation cohort confirmed our initial results and further emphasize the usefulness of the sVNT for rapid screening of larger number of samples for the presence of neutralizing anti-SARS-CoV-2-S-RBD antibodies.

## Discussion

A detailed understanding of immune responses following SARS-CoV-2 infection will enable better treatment and diagnostic procedures, as well as the development of successful vaccines that would help to control the global COVID-19 pandemic. In this regard it is important to gain a better understanding about the presence of neutralizing anti-SARS-CoV-2 serum antibodies in the population as they potentially prevent (re-)infection and even could serve as a treatment option (*31, 32*). Here, we developed a surrogate virus neutralization test that is based on ELISA technology and thus can be adapted to allow for high throughput analysis of samples. We validated this assay by comparing data from the sVNT with that derived from a classical pVNT and found a strong correlation between the results obtained with these two tests. These findings are in line with a pre-print that was made available while preparing this manuscript (*33*). Compared to the pVNT, sVNT is technically less complicated, cheaper and much faster, making it more suitable for a rapid screening of large number of samples. Standardization of this test might, therefore, be potentially important for selection of convalescent plasma donors for the treatment of severe COVID-19 patients. Likewise, since this assay does not rely on anti-species antibodies it can also be applied to detect neutralizing antibodies in any animal species used for pre-clinical testing of SARS-CoV-2 vaccines.

Applying pVNT and sVNT, we found that approximately 90% of recovered COVID-19 patients with mild disease possessed neutralizing serum antibodies. These findings are in line with an early report suggesting that recovered COVID-19 patients have neutralizing SARS-CoV-2-S RBD antibodies in serum after discharge from the hospital (*16*), while other preliminary data indicate that neutralizing SARS-CoV-2-S-RBD antibodies are undetectable in one third of convalescent COVID-19 patients (*29, 34*). Additional studies are therefore required to provide more detailed insight into the levels of neutralizing SARS-CoV-2-S antibodies in convalescent COVID-19 individuals from different countries. Those studies should also exclude false-positive PCR or COVID-19 misdiagnosis as the possible reason for lack of antibodies in certain suspected COVID-19 patients. This would be particularly important as convalescent COVID-19 individuals without neutralizing antibodies might still be susceptible to re-infection and would not be able to provide plasma for the prevention and treatment of COVID-19.

Our data also corroborate other studies indicating that levels of neutralizing antibodies in convalescent COVID-19 individuals are generally low (*29, 34*). Interestingly, a recent pre-print reported that individual neutralizing antibodies against SARS-CoV-2-S-RBD have a half-maximal inhibitory concentration against authentic SARS-CoV-2 ranging from 3 to 709 ng/ml (*29*). Together these data suggest that a considerable proportion of COVID-19 patients with mild disease can mount intermediate to high-affinity IgG antibodies. Since such antibodies are not present in a certain proportion (5% to 30%) of COVID-19 cases, these findings indicate that effective clearance of virus does not rely exclusively on the humoral but also includes cellular immune responses (*12, 14–17*).

The present investigation was done primarily on young and middle-aged convalescent COVID-19 patients with mild disease fulfilling all criteria for blood donation. It is therefore not surprising that in the present study patients’ age did not affect neutralizing anti-SARS-CoV-2-S IgG titers that had been reported by others (*29, 34*). Our data rather indicate that serum levels of total and neutralizing anti-SARS-CoV-2-S antibodies positively correlate with the duration of symptoms, which is also in line with observations made by others (*34*). This hypothesis is further supported by findings that significantly higher titers of neutralizing antibodies were present in sera of severely affected than of the mildly affected patients. However, as another study did not find a correlation between the duration of symptoms and neutralizing anti-SARS-CoV-2-S IgG titers (*29*), it therefore remains to be discerned in future studies how age, gender or disease duration affect the neutralizing antibodies titers.

Altogether, this study reports a high throughput surrogate virus neutralization assay for SARS-CoV-2. Data from this assay highly correlate with results obtained by classical, but laborious and time consuming pseudotyped virus neutralization assay. Both assays revealed the presence of neutralizing anti-SARS-CoV-2-S antibodies, albeit at low titers, in sera of many but not all convalescent COVID-19 patients with mild disease. These findings have implications for selection of convalescent donors for passive immunization by plasma therapy, for which it would be crucial to select donors with relatively high neutralizing antibody titers. It would be also important to address whether any success of passive immunization by plasma therapy would correlate to the total or neutralizing antibody levels in donor’s sera. Furthermore, additional studies are required to understand why neutralizing anti-SARS-CoV-2-S antibodies do not develop in all patients and for how long neutralizing antibodies are present in patients recovered from COVID-19.

## Material and Methods

### Serum samples

Serum samples were collected from convalescent COVID-19 individuals that volunteered for donating plasma at Hannover Medical School’s (HMS) Institute of Transfusion Medicine and Transplant Engineering. All donors had PCR-diagnosed SARS-CoV-2 infection and showed only mild clinical symptoms. Serum was also collected from in-patients with severe COVID-19 symptoms and from healthy controls without any COVID-19-related symptoms (Table S1, S2 and S3). Blood donors consented prior to blood donation and in-patients at time of hospital admission that samples can be used for research purposes. Written informed consent was obtained from all participants. Studies investigating serum samples from healthy controls and COVID-19 patients were approved by the HMS institutional review board (#9001_BO_K2020 and #7901_BO_K2018).

### ELISA

Serum samples were analyzed in the Clinical Virology Laboratory and the Clinic for Rheumatology und Immunology of HMS using the CE-certified versions of the Euroimmun SARS-CoV-2 S1 IgG and IgA ELISA (Euroimmun, Lübeck, Germany) according to the manufacturer’s recommendations.

### Pseudotyped virus neutralization assay

A pseudotyped virus neutralization test (pVNT) based on SARS-CoV-2 protein pseudotyped vesicular stomatitis virus (VSV) particles was performed at HMS’s Institute of Virology and the Primate Center in Göttingen as described earlier (*8*). In brief, pseudotyped VSV particles were produced by calcium-phosphate transfecting HEK293T cells with expression plasmids for the respective glycoproteins, either pCAGGS-VSV-G (*35*) for expression of VSV-G of the control virus or pCG1-SARS-2-SΔ18 (*36*) for the SARS-CoV2 spike protein. 18 hours later the cells were infected with VSV*ΔG-FLuc, a replication deficient recombinant VSV in which the VSV-G open reading frame has been replaced by combined GFP and firefly luciferase expression cassettes (*37*). This VSV*ΔG-FLuc stock virus was propagated in BHK-21 G43 cells (*38*). After incubating the transduced cells with the viral particles for 2 hours at 37°C the supernatant was removed and the cells were washed twice with PBS. Cells were than supplied with medium containing a mAb targeting VSV-G (supernatant from mouse hybridoma CRL-2700; ATCC) to neutralize residual VSV-G, a step that was omitted for the cells transfected with the VSV-G expression construct. 20 hours later the pseudotype particle containing supernatant was separated from the cells by centrifugation and used for the neutralization assays.

For the pVNT Vero76 cells were seeded at 1 × 10^4^ cells per well in 96-well plates. The next day the complement in test sera was inactivated by heating samples to 56°C for 30 min. Sera were then serially diluted, mixed 1:2 with the pseudotyped VSV and incubated for 30 minutes at 37°C. Medium was removed from Vero76 cells and replaced in triplicate wells with the serum/pseudotype particle mixture. 20 hours post infection the supernatant was removed from the cells and replaced with 1x luciferase lysis buffer (2x Lysis juice, 102517, PJK). Cells were lysed for 30 minutes at room temperature. Lysates were transferred to white plates with luciferase substrate (Beetle juice, 102511, PJK) and luciferase activity was measured with a Hidex Sense plate luminometer (Hidex) or a GloMax Discover Microplate Reader (Promega). Data was plotted after background subtraction and normalization to the “no serum” controls. Pseudotyped virus neutralizing titers 50 and 90 (pVNT_50/90_) were defined as the last serum dilutions that reduced transduction efficiency of biological triplicates by at least 50% or 90 % respectively.

### Expression and purification of recombinant soluble ACE2-IgG1 protein

HEK293T cells were grown in DMEM/10% ultra-low IgG FBS/PenStrep and transfected transiently with plasmid pcDNA3-sACE2(WT)-Fc (a gift from Erik Procko; Addgene plasmid # 145163(*39*)) by applying standard calcium phosphate procedures. Supernatants were collected and run over a protein A-sepharose column (ThermoFisher). Bound recombinant protein was eluted with 0.1M sodium citrate pH 3.5. Buffer was exchanged to PBSd and integrity as well as purity were confirmed by analyzing 2 µg of protein on a 10% SDS polyacrylamide gel.

### Surrogate virus neutralization assay

The surrogate virus neutralization assay was developed based on the hypothesis that virus neutralizing antibodies should also interfere with the binding of the RBD of SARS-CoV-2 (SARS-CoV-2-S-RBD) to soluble, surface-immobilized ACE2. To establish this assay hACE2 protein (Trenzyme or in-house produced) was coated at different concentrations in 100 mM carbonate-bicarbonate coating buffer (pH 9.6) on F96-Maxisorp Nunc-Immuno plates (ThermoScientific) at +4°C overnight. After washing in phosphate buffered saline (PBS) with 0.05% Tween 20 (PBST), plates were blocked with 2% bovine serum albumin (BSA, Sigma) and 0.1% Tween 20 in 1x PBS for 1.5 hrs at 37°C. Then, His-tag-conjugated SARS-CoV-2-S-RBD (Trenzyme) was added at different concentrations in carrier buffer (1% BSA and 0.05% Tween in 1x PBS) for 1h at 37°C. Unbound SARS-CoV-2-S-RBD was removed by four PBST washes before anti-His peroxidase-labelled monoclonal antibody (mAb; Clone 3D5, prepared in house) in carrier buffer was added for 1h at 37°C. After final washing, the colorimetric signal was developed by adding chromogenic substrate, 3,3’,5,5’-tetramethylbenzidine (TMB; Sigma) and stopped by adding an equal volume of stop solution (0.2M H_2_SO_4_). Finally, the absorbance readings at 450 nm and 570 nm were acquired using the SpectraMax ID3 microplate reader (Molecular Devices). The K_D_ values of the SARS-CoV-2 binding affinity to ACE2 was calculated from binding curves based on their global fit using one-site specific binding analysis (GraphPad Prism).

To test for the presence of neutralizing anti-SARS-CoV-2-S serum antibodies, 6 ng of SARS-CoV-2 S RBD was pre-incubated with test sera at final dilutions between 1:20 to 1:540 as indicated on the graphs for 1 h at 37°C, before adding them to plates coated with 150 or 300 ng/well ACE2. For each reaction, the percentage of inhibition was calculated from optical density values after subtraction of background values as: Inhibition (%) = (1 - Sample OD value/Average SARS-CoV-2 S RBD OD value) x100. Neutralizing sVNT titers were determined as dilution that still had binding reduction > mean + 2 SD of values from sera of healthy controls.

### Statistical analysis

Linear data were analyzed using unpaired t-test with Welch’s correction (for 2 groups) or Welch’s ANOVA followed by Dunnett’s T3 multiple comparisons test (for 3 groups) and were correlated using Pearson r test. Categorical data were analyzed using χ^2^ test or Fisher’s exact test for unpaired proportions, as indicated beneath each figure. Correlation between linear and categorical data was done using ordinary one-way ANOVA followed by test for linear trend. All statistical analyses were conducted using GraphPad Prism 8.4 (GraphPad Software, USA).

## Data Availability

All data are available upon request to the corresponding authors.

## H2:Supplementary Materials

## General

We thank all study participants for providing us with the samples.

## Funding

This work was supported by Deutsche Forschungsgemeinschaft, DFG Excellence Strategy – EXC 2155”RESIST”–Project ID39087428 and by funds of the state of Lower Saxony (14-76103-184 CORONA-11/20) to RF and (1476103-184 CORONA-12/20) to TFS and by funds of BMBF (RAPID consortium, 01K11723D).

## Author contributions

RF conceived and guided the study. RF and BB developed sVNT. BB, SCS, AKC, GB, IR, WP, AC, SW and CR performed laboratory work including assay setup and data analysis. SP, MH and HK-W initially developed and performed and TFS coordinated conventional neutralization assays at HMS. RB, NG, MY, IP, and JM set up the Hanover convalescent donor registry, coordinated donor testing and provided samples. CRS-F analyzed clinical patient data and provided healthy control samples.

GMNB, IP, MMH, and TFS also provided samples. BB and RF wrote the manuscript with input from all authors.

## Competing interests

We declare no competing interests.

---

**Fig S1.**
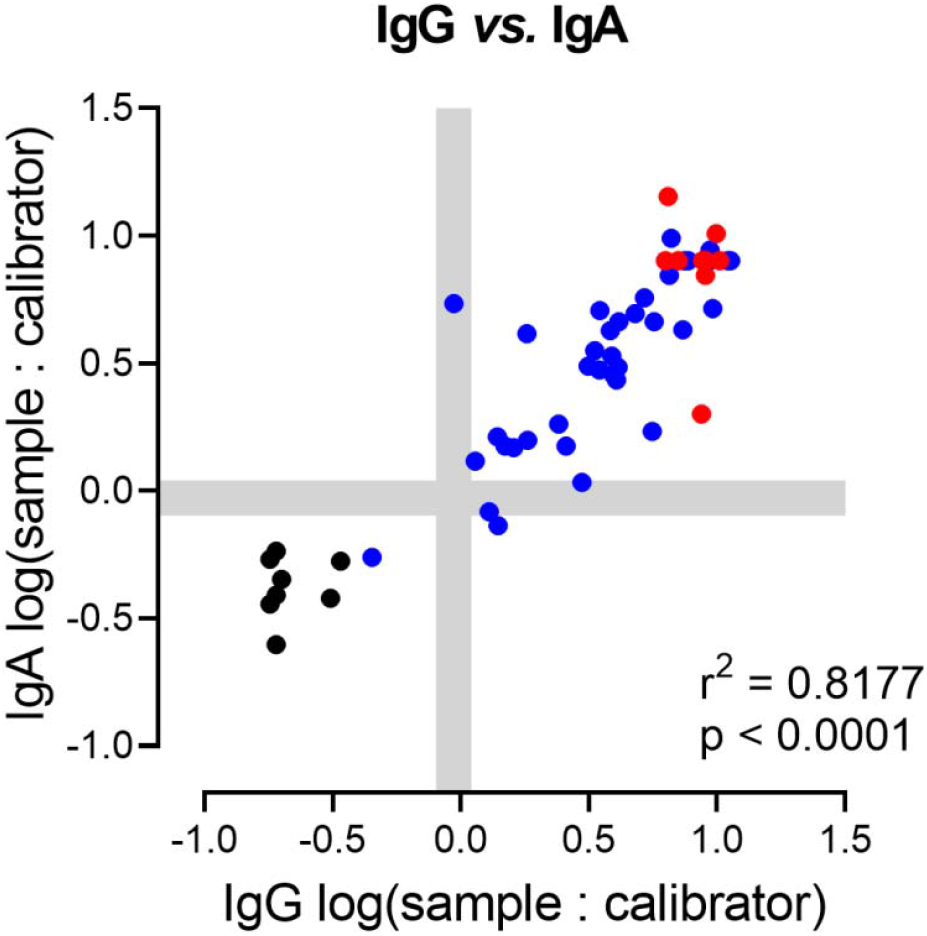
Total IgG and IgA antibody levels against SARS-CoV-2 strongly correlate. Samples from mild (blue) and severe (red) COVID-19 convalescent patients and HC (black dots). Shaded areas, vendor-defined cut-off values to determine positive, borderline and negative samples as described for figure 1. Correlation, Pearson r.

**Fig S2.**
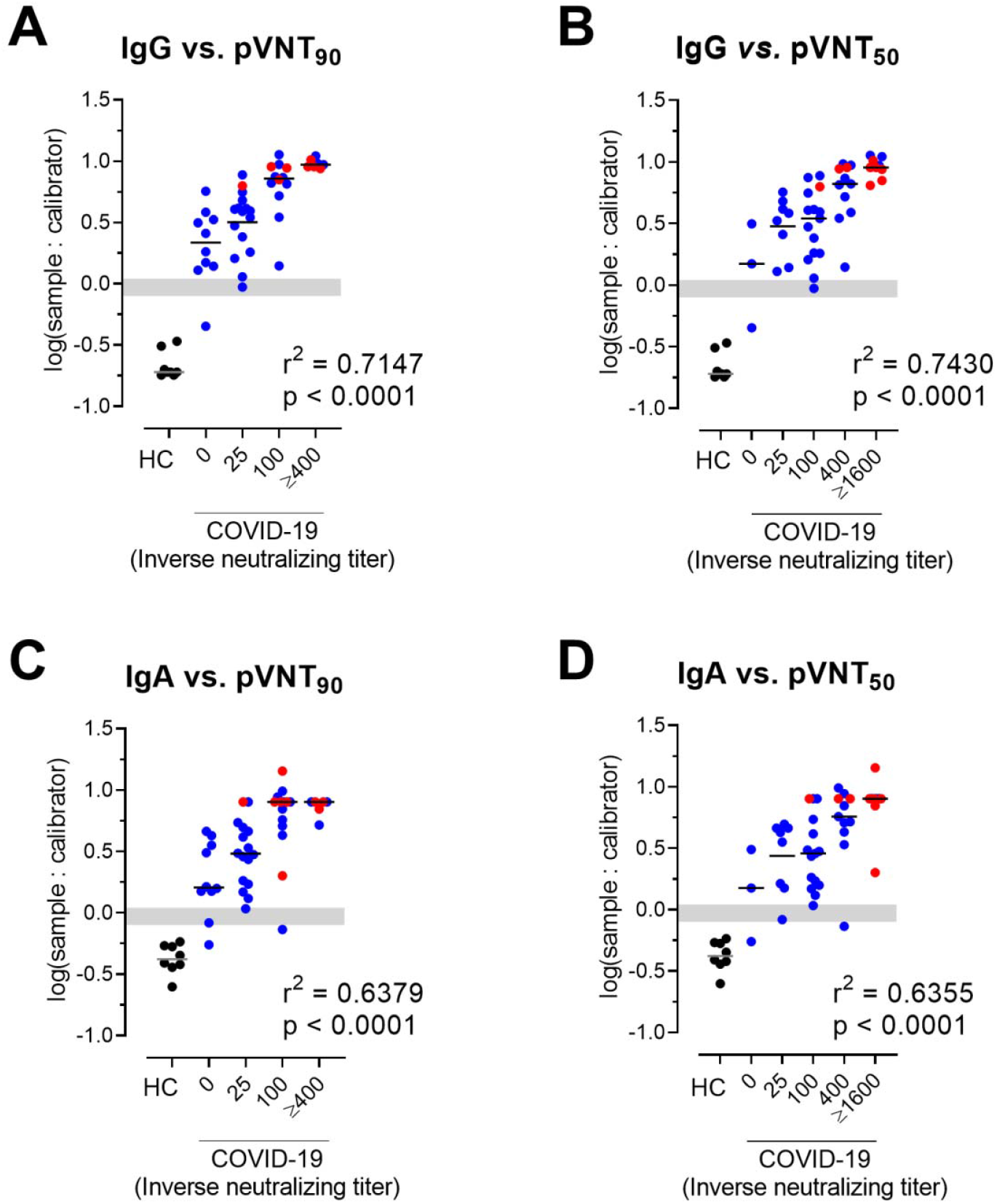
Positive correlation between pVNT and anti SARS-CoV-2-S1 IgG and IgA antibodies (A-D) Anti SARS-CoV-2-S1-specific IgG and IGA levels determined by ELISA and pVNT_90_ or pVNT_50_ neutralizing antibody titers as indicated. Dots, samples of HC (black), mildly (blue) or severely (red) affected COVID-19 convalescent patients. Shaded areas indicate cut-off values to determine positive (above), borderline (within) and negative (below gray area) samples. Correlation, one-way ANOVA followed by test for trend.

**Fig S3.**
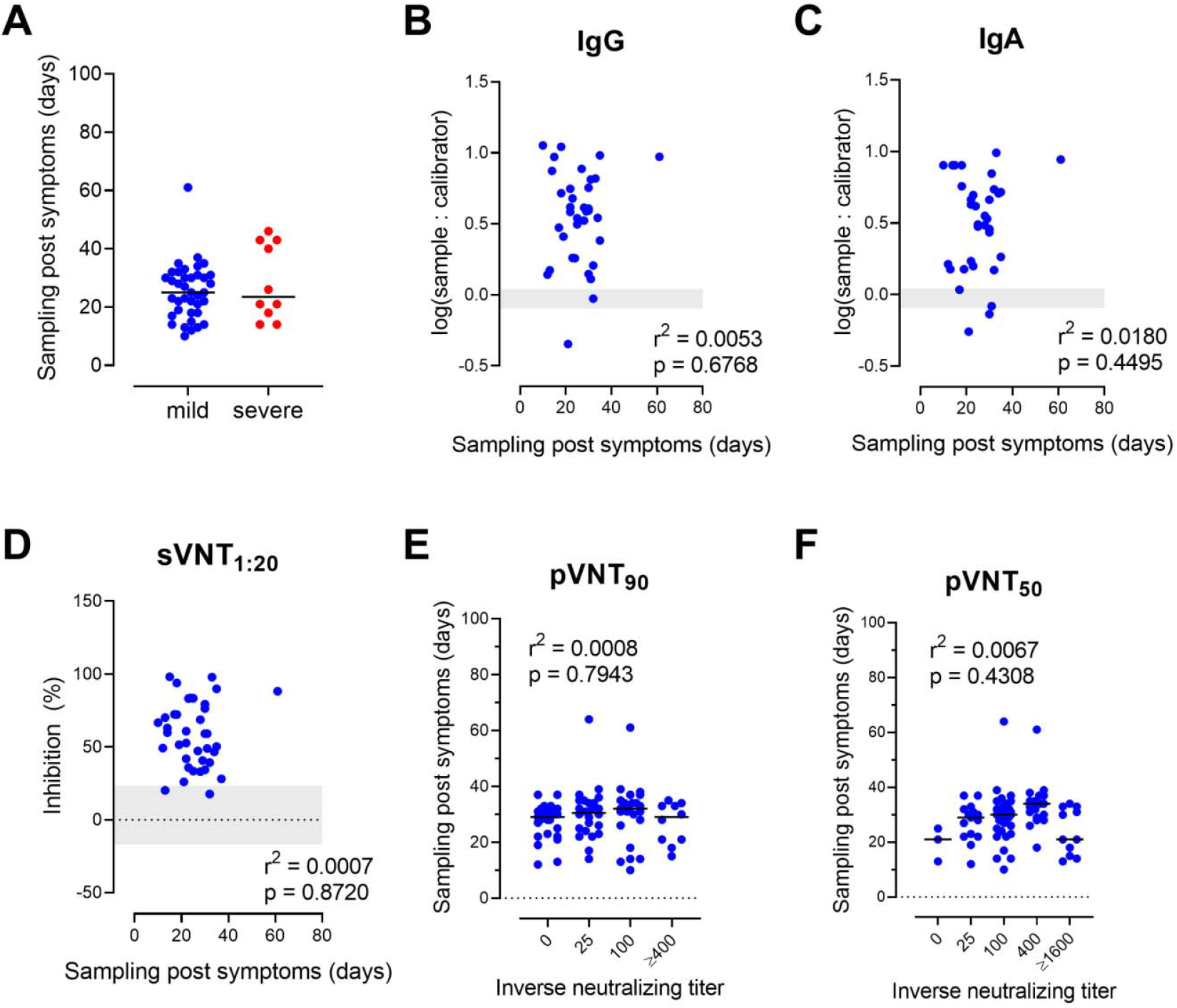
No correlation between time of sampling and total and neutralizing SARS-CoV-2-specific antibody levels. (A) No difference between date of sampling between convalescent patients with mild and severe COVID-19. Dots, individuals; bars, mean, * P < 0.05, Welch’s t test. (B-F) No correlation between time of sampling (post occurrence of symptoms) and log-transformed SARS-CoV-2-S1-specific IgG (B) and IgA (C) levels, sVNT_1:20_ (D), pVNT_90_ (E) or pVNT_50_ (F) in convalescent patients with mild COVID-19. Dots, individuals, (E, F) horizontal lines, medians. (B,C) Shaded areas indicate vendor-defined cut-off values to determine positive (above), borderline (within) and negative (below gray area) samples. (D) The shaded area indicates mean ± 2 SD range of inhibition of sera from HC; Correlation, Pearson r (B-D) or one-way ANOVA followed by test for trend (E,F).

**Fig S4.**
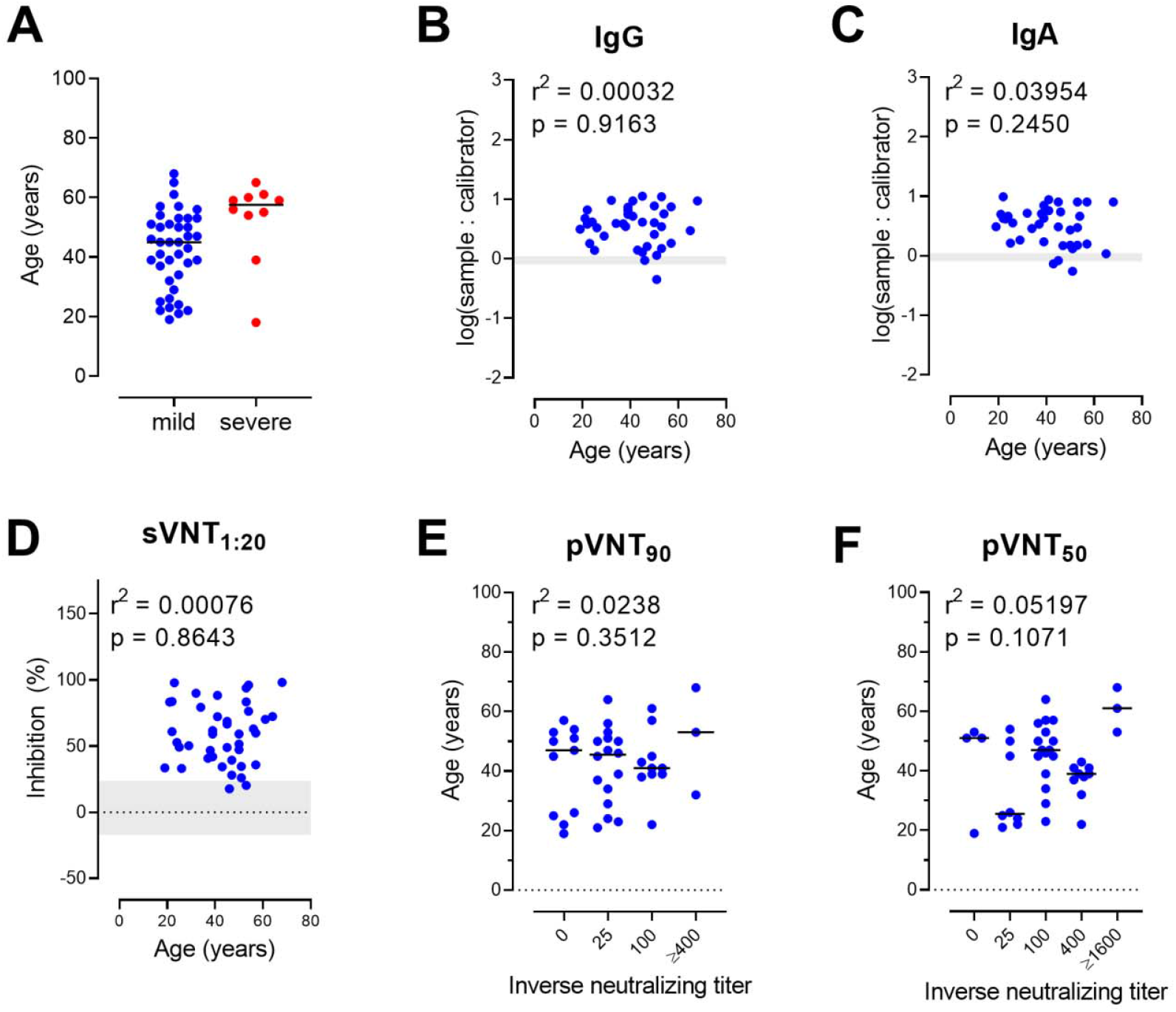
No correlation between patient age and total and neutralizing SARS-CoV-2-S-specific antibody levels. (A) Convalescent patients with severe COVID-19 were significantly older than patients with mild disease. Dots, individuals; bars, mean, * P < 0.05, Welch’s t test. (B-F) No correlation between age and log-transformed SARS-CoV-2-S-specific IgG (B) and IgA (C) levels, sVNT_1:20_ (D), pVNT_90_ (E) or pVNT_50_ (F). Dots, individuals recovered from mild COVID-19 (E, F) horizontal lines, medians. (B,C) Shaded areas indicate vendor-defined cut-off values to determine positive (above), borderline (within) and negative (below gray area) samples. (D) The shaded area indicates mean ± 2 SD range of inhibition of sera from HC; Correlation, Pearson r (B-D) or one-way ANOVA followed by test for trend (E,F).

**Fig S5.**
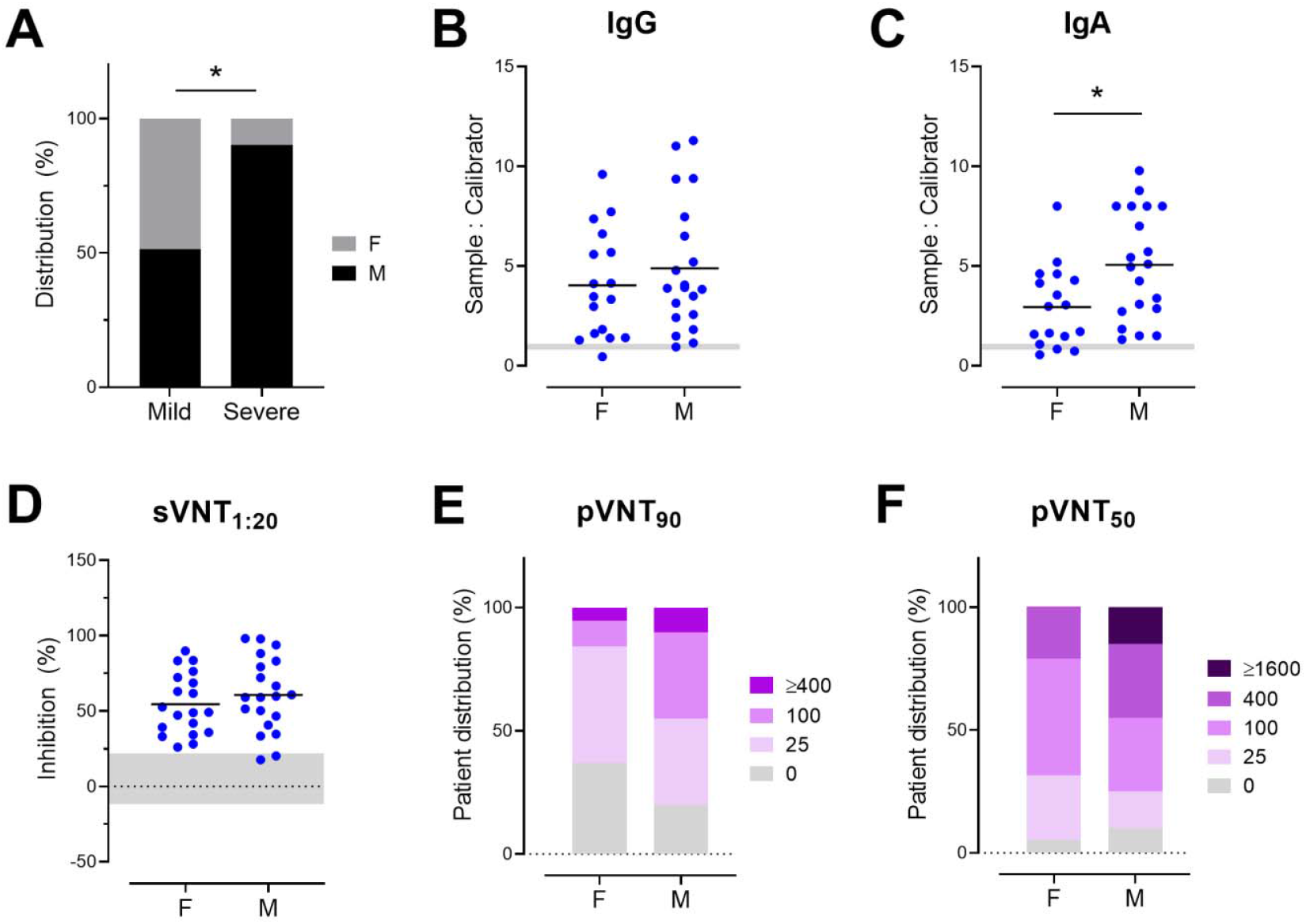
Male convalescent patients suffered more frequently than females from severe COVID-19 and produce higher anti-SARS-CoV-2 S IgA antibody levels. (A) Gender distribution in patients recovered from mild or severe COVID-19. * P < 0.05, Fisher’s exact test. (B-F) Female (F) and male (M) convalescent patients with mild COVID-19 have similar levels of serum anti-S1 IgG antibodies (A) while IgA levels are significantly higher in males (B). No differences in sVNT_1:20_ (C) or sVNT (D), pVNT_90_ (E) or pVNT_50_ (F) between female and male COVID-19 convalescent individuals with mild disease. (B-D) Dots, individuals; horizontal bars, means. (B,C) Shaded areas, vendor-defined ELISA cut-off values that define positive (above shaded area), borderline (within shaded area) and negative (below shaded area) samples. (E) Shaded area, mean ± 2 S.D. of inhibition of sera from HC. (B-D) unpaired t-test; (E-F) Chi-square test; * P < 0.05.

**Fig s6.**
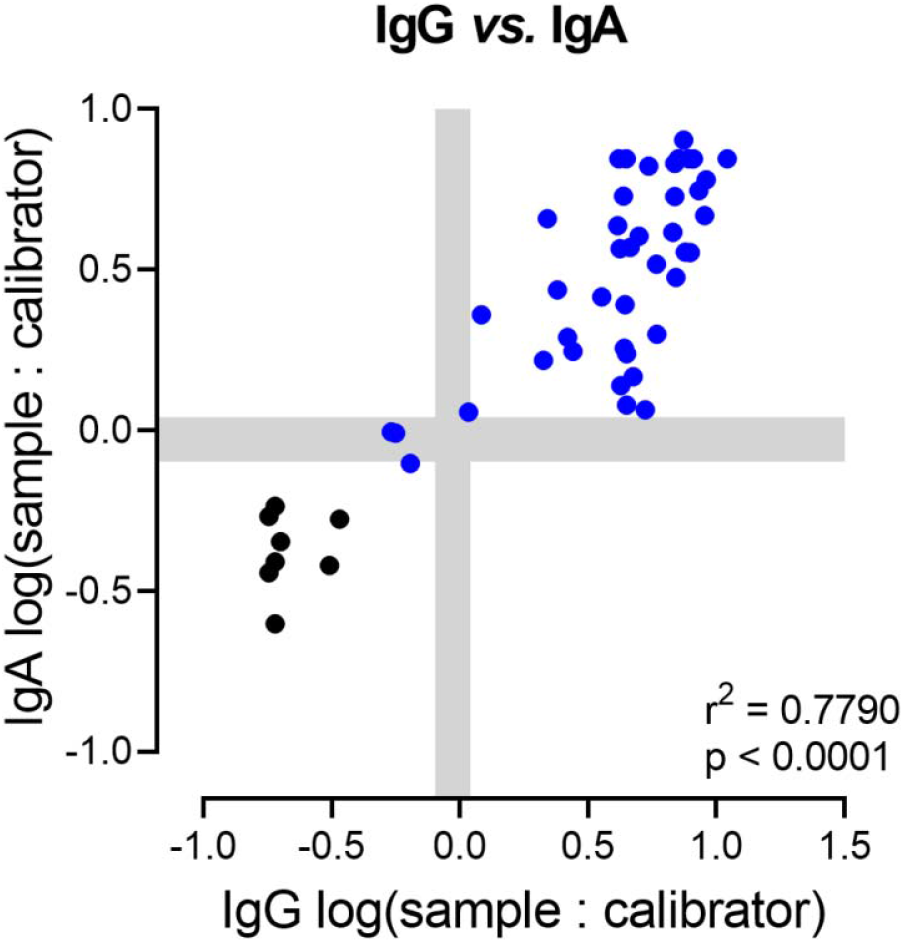
Total IgG and IgA antibody levels against SARS-CoV-2-S1 strongly correlate in the validation group. Samples from mild (blue) COVID-19 convalescent patients and the initial 12 HC (black dots). Shaded areas, vendor-defined cut-off values to determine positive, borderline and negative samples as described for figure 1. Correlation, Pearson r.

**Fig S7.**
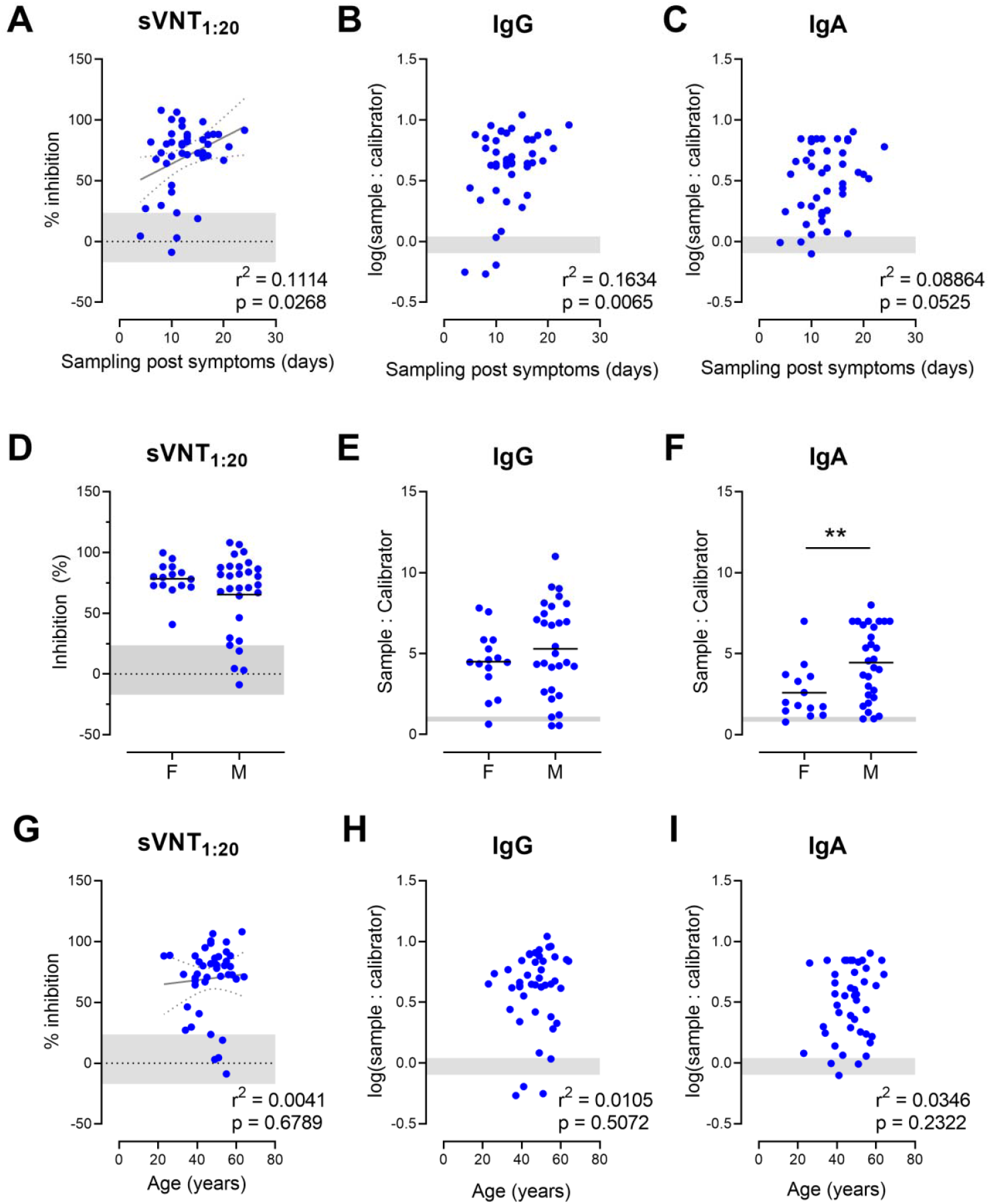
Duration of symptoms but not patient age or gender positively correlates to total and neutralizing SARS-CoV-2-S-specific antibody levels in the validation group of convalescent patients with mild COVID-19 (A-C) Weak positive correlation between duration of symptoms and sVNT_1:20_ (A) and log-transformed SARS-CoV-2-S1-specific IgG (B) but not IgA (C) levels. (D-F) No difference between female (F) and male (M) COVID-19 convalescent individuals with mild disease regarding sVNT_1:20_ (D) and serum levels of SARS-CoV-2-S1-specific IgG antibodies (E) while levels of serum SARS-CoV-2-S1-specific IgA antibodies are significantly higher in males (F). ** P < 0.01, Welch’s t test. (G-I) No correlation between age of patients recovered from mild COVID-19 and sVNT_1:20_ (G) or log-transformed anti SARS-CoV-2-S1-specific IgG (H) and IgA (I) antibody levels Dots, individuals; horizontal lines, means. Shaded areas indicate mean ± 2 SD of inhibition of sera from HC (A,D,G) or the respective cut-off values from ELISA as described for Fig 5 (B,C,E,F,H,I). Correlation, Pearson r (A-C,G-I); Comparison Welch’s t test (D-F).

**Table S1.**
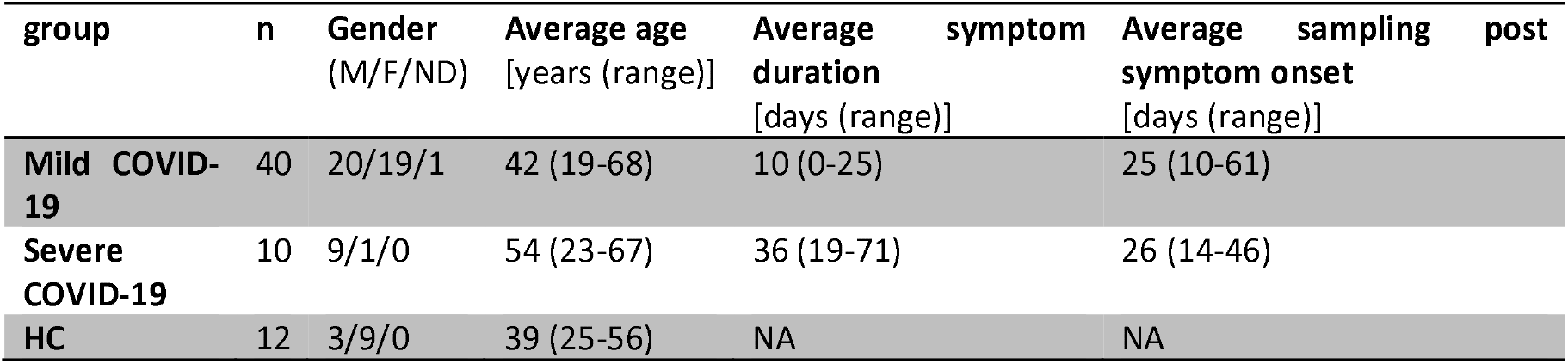
Cohort characteristics. Abbreviations: HC – healthy controls; M –male; F-female; NA – not applicable; ND – not disclosed.

**Table S2.** Demographic and clinical characteristics of individuals with mild and severe COVID-19 and healthy controls (HC). Abbreviations: PEMC - pre-existing medical conditions; m: male; f: female; NA: not available; *: in days; §: days after onset of symptoms.

| ID | Gender | Age | PEMC | COVID-19 | Symptoms | ECMO or intubation | Symptom duration* | Sampling§ |
| --- | --- | --- | --- | --- | --- | --- | --- | --- |
| M-01 | F | 20-25 | no | mild | NA | no | 5 | 12 |
| M-02 | M | 45-50 | no | mild | Fatigue, cough, ageusia, body aches, headache | no | 14 | 10 |
| M-03 | M | 50-55 | no | mild | NA | no | NA | NA |
| M-04 | NA | 60-65 | no | mild | Limb pain | no | 18 | 13 |
| M-05 | M | 65-70 | yes | mild | Fever, chills, cough, body aches, sore throat, | no | 20 | 15 |
| M-06 | f | 65-70 | no | mild | Fever, Sore throat, diarrhea (once) | no | 13 | 17 |
| M-07 | M | 20-25 | no | mild | No symptoms | no | 0 | 23 |
| M-08 | F | 55-60 | no | mild | Cough, chills | no | 3 | 23 |
| M-09 | M | 55-60 | yes | mild | Cough, fever, headache, body aches | no | 18 | 14 |
| M-10 | F | 55-60 | no | mild | Chills, fever, diarrhea | no | 19 | 14 |
| M-11 | F | 20-25 | no | mild | Cough, headache, body aches, swollen lymph nodes (4 days) | no | 9 | 22 |
| M-12 | M | 40-45 | no | mild | fever, cough | no | 10 | 18 |
| M-13 | M | 50-55 | no | mild | Fatigue, cough, headache, anosmia, body aches | no | 16 | 18 |
| M-14 | F | 50-55 | no | mild | Cough, anosmia, rhinitis, ageusia | no | 10 | 21 |
| M-15 | F | 35-40 | no | mild | Mild cough | no | NA | NA |
| M-16 | F | 50-55 | no | mild | Body ache (2 days), loss of smell | no | 10 | 27 |
| M-17 | M | 25-30 | no | mild | Rhinitis | no | 5 | 35 |
| M-18 | M | 50-55 | yes | mild | Cough, fever | no | 25 | 13 |
| M-19 | F | 50-55 | no | mild | Headache, shortness of breath, pressure on the bronchi | no | 10 | 30 |
| M-20 | M | 15-20 | no | mild | Cough, fever | no | 9 | 25 |
| M-21 | M | 50-55 | no | mild | Fever, headache, lung pain | no | 10 | 19 |
| M-22 | M | 20-25 | no | mild | Fatigue, cough | no | 12 | 22 |
| M-23 | F | 40-45 | no | mild | Light symptoms | no | 7 | 31 |
| M-24 | F | 25-30 | no | mild | Mild fever, cough, fatigue, chills | no | 9 | 28 |
| M-25 | M | 35-40 | no | mild | Cough, sore throat, fever | no | 8 | 31 |
| M-26 | M | 50-55 | no | mild | Diarrhea, cough, rhinitis, fever | no | 9 | 30 |
| M-27 | F | 35-40 | no | mild | Fever, headache, fatigue | no | 14 | 22 |
| **M-28** | M | 45-50 | no | mild | Cough | no | 1 | 32 |
| **M-29** | M | 35-40 | no | mild | Fever, body aches, anosmia, ageusia | no | 5 | 34 |
| **M-30** | F | 45-50 | no | mild | Headache, fatigue | no | 1 | 37 |
| **M-31** | F | 40-45 | yes | mild | Cough, headache, dyspnea | no | 6 | 30 |
| **M-32** | F | 40-45 | yes | mild | Cough, fatigue | no | 8 | 28 |
| **M-33** | F | 45-50 | no | mild | Fever, light cough, bedridden for 2 days | no | 5 | 32 |
| **M-34** | M | 35-40 | no | mild | Fever, chills, fatigue, cough, running nose | no | 13 | 29 |
| **M-35** | M | 30-35 | no | mild | Sore throat, elevated temperature, cough, fatigue | no | 4 | 30 |
| **M-36** | M | 20-25 | no | mild | Fever, sore throat | no | 5 | 33 |
| **M-37** | F | 20-25 | no | mild | Fever, body aches, rhinitis, sore throat | no | 4 | 24 |
| **M-38** | F | 50-55 | no | mild | Fever, dyspnea, headache | no | 11 | 25 |
| **M-39** | M | 40-45 | NA | mild | Body aches, ageusia, chills | no | 14 | 61 |
| **M-40** | F | 30-35 | no | mild | Fever, diarrhea, body aches, headache, pneumonia, cough | no | 16 | 35 |
| **S-01** | F | 50-55 | no | severe | Fever, body aches, dyspnea, dizziness | intubation | 28 | 21 |
| **S-02** | M | 55-60 | NA | severe | Dyspnea, fever | intubation | 35 | 21 |
| **S-03** | M | 60-65 | yes | severe | Fever, dyspnea, pneumonia, cough | intubation | 30 | 26 |
| **S-04** | M | 50-55 | NA | severe | Fever, dyspnea, body aches, ageusia, chills | intubation | 29 | 40 |
| **S-05** | M | 60-65 | no | severe | Fever, diarrhea, dry cough | ECMO | 71 | 43 |
| **S-06** | M | 50-60 | yes | severe | Dyspnea, fever, dry cough | intubation | 67 | 46 |
| **S-07** | M | 35-40 | no | severe | Dyspnea, fever | intubation | 55 | 43 |
| **S-08** | M | 65-70 | yes | severe | Dyspnea, fever | ECMO | 28 | 14 |
| **S-09** | M | 55-60 | yes | severe | Dyspnea, fever | intubation | 19 | 14 |
| **S-10** | M | 15-20 | no | severe | Loss of smell, fever, nausea, emesis, dyspnea | no | 21 | 18 |
| **S-01** | F | 50-55 | no | severe | Fever, body aches, dyspnea, dizziness | intubation | 28 | 21 |
| **HC-01** | F | 35-40 | NA | NA | NA | NA | NA | NA |
| **HC-02** | F | 30-35 | NA | NA | NA | NA | NA | NA |
| **HC-03** | F | 35-40 | NA | NA | NA | NA | NA | NA |
| **HC-04** | F | 20-25 | NA | NA | NA | NA | NA | NA |
| **HC-05** | F | 50-55 | NA | NA | NA | NA | NA | NA |
| **HC-06** | F | 45-50 | NA | NA | NA | NA | NA | NA |
| **HC-07** | M | 20-25 | NA | NA | NA | NA | NA | NA |
| HC-08 | M | 45-50 | NA | NA | NA | NA | NA | NA |
| HC-09 | F | 55-60 | NA | NA | NA | NA | NA | NA |
| HC-10 | F | 30-35 | NA | NA | NA | NA | NA | NA |
| HC-11 | F | 50-55 | NA | NA | NA | NA | NA | NA |
| HC-12 | M | 30-35 | NA | NA | NA | NA | NA | NA |

**Table S3.** Clinical characteristics of confirmation mild COVID-19 cases.

| group | n | Gender (M/F/ND) | Average age<br>[years (range)] | Average symptom duration<br>[days (range)] | Average sampling post symptom onset<br>[days (range)] |
| --- | --- | --- | --- | --- | --- |
| Confirmation mild COVID-19 | 44 | 29/15/0 | 47 (23-64) | 13 (4-24) | 33 (14-64) |

| ID | Gender | Age | PEMC | COVID-19 | Symptoms | ECMO or intubation | Symptom duration* | Sampling§ |
| --- | --- | --- | --- | --- | --- | --- | --- | --- |
| M-41 | M | 60-65 | no | mild | fever, diarrhea, anosmia, ageusia | no | 8 | 34 |
| M-42 | F | 40-45 | yes | mild | headache, body aches, anosmia, ageusia, fever (light), rhinitis | no | 13 | 27 |
| M-43 | M | 35-40 | NA | mild | dyspnea, fever, headache, sore throat | no | 10 | 33 |
| M-44 | F | 55-60 | yes | mild | cough, rhinitis, slightly elevated temperature | no | 12 | 28 |
| M-45 | M | 45-50 | no | mild | cough, fatigue (5 days), pressure on the lungs | no | 11 | 33 |
| M-46 | M | 35-40 | no | mild | fever, headache, diarrhea, chill, cough, anosmia, ageusia | no | 8 | 30 |
| M-47 | M | 55-60 | no | mild | cough (light), rhinitis | no | 16 | 31 |
| M-48 | M | 25-30 | no | mild | rhinitis, anosmia, ageusia | no | 10 | 31 |
| M-49 | M | 45-50 | no | mild | fever, joint pain, bitter taste, pneumonia, headaches | no | 10 | 26 |
| M-50 | M | 50-55 | no | mild | cough (light), tiredness, loss of taste | no | 9 | 33 |
| M-51 | F | 45-50 | no | mild | increased temperature, snuff (light), loss of taste | no | 6 | 39 |
| M-52 | M | 40-45 | yes | mild | cough (light), aching limbs, headaches | no | 16 | 34 |
| M-53 | M | 40-45 | yes | mild | fever, aching limbs, loss of smell and taste, cough | no | 20 | 28 |
| M-54 | F | 40-45 | no | mild | headaches, cough, weld outbreaks, fever | no | 17 | 36 |
| M-55 | F | 45-50 | yes | mild | aching limbs, respiratory problems, loss of smell, fever, cough, fatigue | no | 21 | 34 |
| M-56 | M | 45-50 | no | mild | fever, fatigue, loss of smell, passed out (one time), diarrhoea | no | 11 | 37 |
| M-57 | M | 45-50 | yes | mild | fever (light), cough (light), aching limbs | no | 13 | 37 |
| M-58 | M | 45-50 | no | mild | fever, sore throat, headaches, fatigue, loss of smell and taste | no | 17 | 30 |
| M-59 | F | 55-60 | yes | mild | cough, nausea, aching limbs, restricted sense of smell | no | 12 | 37 |
| M-60 | M | 60-65 | no | mild | fever | no | 16 | 38 |
| **M-61** | M | 45-50 | no | mild | fever, aching limbs, loss of smell and taste, cough | no | 13 | 35 |
| **M-62** | F | 40-45 | no | mild | fever, aching limbs, cough | no | 12 | 39 |
| **M-63** | M | 55-60 | no | mild | cough drier, fever, snuff, shivering, headaches, loss of smell and taste, severe fatigue, light bronchitis | no | 24 | 37 |
| **M-64** | M | 50-55 | no | mild | loss of smell and taste, diarrhoea, dry cough, loss of appetite | no | 15 | 34 |
| **M-65** | M | 35-40 | no | mild | fever (light), fatigue | no | 9 | 64 |
| **M-66** | F | 30-35 | n.a. | mild | keine Symptome eingetragen | no | 8 | 32 |
| **M-67** | M | 35-40 | no | mild | fever, cough, sore throat, shivering, apathy, fatigue | no | 10 | 29 |
| **M-68** | M | 50-55 | no | mild | Cough, fever (1 day), headache, body aches | no | 17 | 26 |
| **M-69** | M | 50-55 | no | mild | fatigue, chills, body aches | no | 12 | 32 |
| **M-70** | F | 55-60 | no | mild | chills, fever (light), headache, fatigue, body aches | no | 15 | 29 |
| **M-71** | M | 45-50 | yes | mild | fever, cough, headache, dyspnea | no | 10 | 30 |
| **M-72** | F | 55-60 | yes | mild | Headache, sore throat, anosmia, ageusia, tinnitus | no | 12 | 34 |
| **M-73** | F | 35-40 | no | mild | conjunctivitis, cough, fever, anosmia, ageusia, diarrhea | no | 19 | 31 |
| **M-74** | F | 60-65 | yes | mild | cough, sore throat, headache, fatigue, vertigo, blood pressure fluctuation | no | 16 | 35 |
| **M-75** | F | 20-25 | no | mild | fever, fatigue, body aches, cold symptoms | no | 13 | 31 |
| **M-76** | F | 50-55 | no | mild | headache, body aches, ageusia | no | 13 | 33 |
| **M-77** | M | 45-50 | yes | mild | fever, fatigue, anosmia, ageusia | no | 16 | 32 |
| **M-78** | M | 55-60 | yes | mild | cough, headache, body aches, fever (light) | no | 18 | 14 |
| **M-79** | F | 40-45 | no | mild | fever, cough, ageusia, rhinitis | no | 10 | 28 |
| **M-80** | M | 35-40 | yes | mild | no symptoms | no | 7 | 31 |
| **M-81** | M | 45-50 | yes | mild | cold symptoms | no | 11 | 22 |
| **M-82** | M | 50-55 | no | mild | cough, ageusia | no | 10 | 32 |
| **M-83** | M | 30-35 | no | mild | slightly elevated temperature, sore throat, cough, fatigue | no | 5 | 32 |
| **M-84** | M | 50-55 | no | mild | ageusia (very briefly), nausea for a few days in the morning | no | 4 | 30 |

## Notes

### Competing Interest Statement

The authors have declared no competing interest.

### Author Declarations

Studies investigating serum samples from healthy controls and COVID-19 patients were approved by the HMS institutional review board (#9001_BO_K2020 and #7901_BO_K2018).

